# Integrating Online Georeferenced Epidemiological Analysis and Visualization into a Telemedicine Infrastructure – First Results

**DOI:** 10.1101/19000554

**Authors:** Aldo von Wangenheim, Alexandre Savaris, Adriano Ferretti Borgatto, Andrei de Souza Inácio

## Abstract

With the objective to perform a first evaluation of the impact of the integration of a graphic spatial epidemiology tool that allows quasi-realtime georeferenced data visualization into a telemedicine infrastructure, this work presents GISTelemed, an online module specialized on indexing structured and semi-structured data, as well as querying the indexed content using structured and free-text search.

We evaluated GISTelemed accordingly to the guidelines published by the US Centers for Disease Control and Prevention and input provided by a questionnaire customized according to AdEQUATE (questionnAire for Evaluation of QUAlity in TElemedicine systems).

39 healthcare professionals from 13 municipalities participated in the evaluation. We analyzed data from questionnaires using descriptive statistics, being Lernability and Comfort the characteristics that received the best evaluation. Quantitative evaluation based upon leprosis cases detected through tele-dermatology showed a sensitivity and PPV of respectively 77.2% and 95.3%. 22.8% of the cases detected were un-notified cases.

Results from our case study show a good evaluation regarding the perceived software quality”. We conclude that the integration of spatial epidemiology tools to the STT/SC system, besides enabling visualization of data in maps, allowed users to analyze the evolution of morbidities and their co-occurrences.

## INTRODUCTION

Data analysis regarding mortality and survival rates has allowed the examination of multiple dimensions of health.[1] Spatial Epidemiology is the description and analysis of geographic variations in disease. In the last years, advances in Geographic Information Systems (GIS) and availability of high-resolution, geographically referenced health and environmental quality data have created unprecedented new opportunities to investigate local geographic variations in disease.[2] In this context, data relevant to the understanding of health and disease are nowadays found in diverse sources besides formal Electronic Health Records (EHR), but also in imaging, teleconsulting and other data, indicating that, for better epidemiological analyses, we need big data approaches that are able to integrate these data also.[3] Additionally, GIS offer unique opportunities to integrate data across multiple databases and spatial scales for display, management, and analysis.[4]

On the other hand, emerging countries have experienced fast and complex epidemiological changes due to the increasing rates of chronic, communicable and noncommunicable diseases. [5-6] In this context, Brazil has been employing information systems developed for its National Unified Public Health System (SUS), including the Brazilian National Information System on Reportable Diseases (SINAN) for collecting and processing data on mandatory communicable diseases.[7]

Event notification processes, however, are sensitive to incomplete and/or incorrect data, resulting in the inaccurate evaluation of epidemiological studies performed on these data.[8] A number of studies have reported difficulties in collecting reliable data for many communicable diseases, compromising its epidemiological monitoring.[9-12] The lack of widespread EHRs and interoperability gaps between systems contribute on delays and on disseminating errors.[9]

To be able to rely additionally on other systems can help support epidemiological monitoring.

As supporting tools for data acquisition and consolidation, telemedicine technologies, which are, by nature, integrated and online, have become relevant as a basis for epidemiological studies.[13-15] Due to its distributed, unconstrained geographical reach, such technologies allow the data acquisition from telediagnosis and teleconsultation, assisting the epidemiological surveillance through online identification and monitoring of public health issues.[16] The acquired data could contribute to improve epidemiological investigations and disease control, helping healthcare policy makers to direct resources and relocate professionals based on concrete scenarios.[17-18] Despite this potential as epidemiological evidence gathering tools, telemedicine systems have not yet been integrated into epidemiological databases or search tools.[2-3]

An example of a well-established telemedicine infrastructure is the Santa Catarina State Integrated Telemedicine and Telehealth System (STT/SC), a statewide public telemedicine network located at the State of Santa Catarina, Brazil. Developed since 2005 through a partnership between the Santa Catarina’s State Health Department (SES/SC) and the Federal University of Santa Catarina (UFSC), STT/SC provides asynchronous telemedicine and telehealth services via web and mobile applications, supporting radiology, electrocardiography, dermatology, clinical analyses, and healthcare education.[19-20] The network stores structured and semi-structured data for more than five million examinations.

## OBJECTIVES AND STATE OF THE ART

The main research question leading this research was: In a country lacking real-time and short-time disease monitoring resources such as Brazil, is it useful to employ the online-readiness and real-time nature of a Telemedicine infrastructure to provide georreferenced epidemiological data?

With the objective to evaluate the integration of a spatial epidemiology tool that allows georeferenced data visualization into a telemedicine infrastructure, this work presents GISTelemed - a module of STT/SC specialized on indexing structured and semi-structured data, as well as querying the indexed content using structured and free-text search predicates. GISTelemed offers quasi-real-time georefrenced data, collected from different sources, ranging from patient interviews at primary care facilities to high-complexity imaging examinations such as MRI and CT, integrated into a GIS. It allows small-area analyses, encompassing disease mapping and geographic correlation studies. This creates new opportunities to investigate local geographic variations in disease [2]. Preliminary results attest its viability, allowing the multiple-source telemedicine content to be used as a basis for epidemiological studying and monitoring.

We evaluate GISTelemed using both the guidelines for evaluating public health surveillance systems, published by the US Centers for Disease Control and Prevention (CDC),[21] and also AdEQUATE (questionnAire for Evaluation of QUAlity in TElemedicine systems), an evaluation model customized to specificities and needs of telemedicine systems based on the ISO/IEC 25010 standard, the Technology Acceptance Model (TAM), and the System Usability Scale (SUS).[22-25]

A detailed literature review on spatial epidemiology systems and tools would exceed the scope of this paper, but an analysis of the state of the art of the late 1990’s and early 2000’s.is given in [2] and a discussion of the present state of the art is given in [26]. We could not identify any database or tool with the search and visualization capabilities of the GISTelemed that encompasses data from a population such as the population of an entire State of a large Latin American country and also updates on quasi real-time.

## MATERIALS AND METHODS

GISTelemed is a module responsible for the extraction, georeferencing, indexing, search, retrieval, and visualization of content acquired from data sources managed by STT/SC.

### Architecture of GISTelemed

The system offers a dataflow organized in five steps, as follows (see figure 1):

- <u>ETL (Extract, Transform, Load) process</u> - responsible for accessing structured (relational database instances) and semi-structured (PACS - Picture Archiving and Communication System instances) data sources, collecting its content for indexing. Individual ETL jobs were created to satisfy specificities of each dataset (e.g. clinical analyses for HIV, dengue fever, hepatitis B, and toxoplasmosis from the Central Laboratory of Public Health - LACEN - information system, examinations in cardiology and dermatology from the STT information system, DICOM - Digital Imaging and Communications in Medicine - objects from PACS), being executed by a multiprotocol, open source interconnection engine (Mirth Connect). The acquired content is processed and converted into XML (eXtensible Markup Language) documents, sent via HTTP (HyperText Transfer Protocol) to the search engine component.
- <u>Search engine component</u> - responsible for indexing content received from the ETL process, as well as for the execution of search and retrieval according to queries sent from the user interface. Built on top of an open source search server (Apache Solr), the search engine uses information retrieval techniques (e.g. stemming, stop words) to improve free text and structured query processing.
- <u>Pre- and post-processing operations</u> - responsible, respectively, for the standardization of input filters selected by users and for the normalization of query results to be send to the GUI (Graphical User Interface).
- <u>User interface</u> - responsible for the interaction of both experienced and inexperienced users with the system, it organizes input filters allowing the configuration of on-demand queries together with the choice of results to be displayed in tables, charts, and maps.

**Figure 1.**
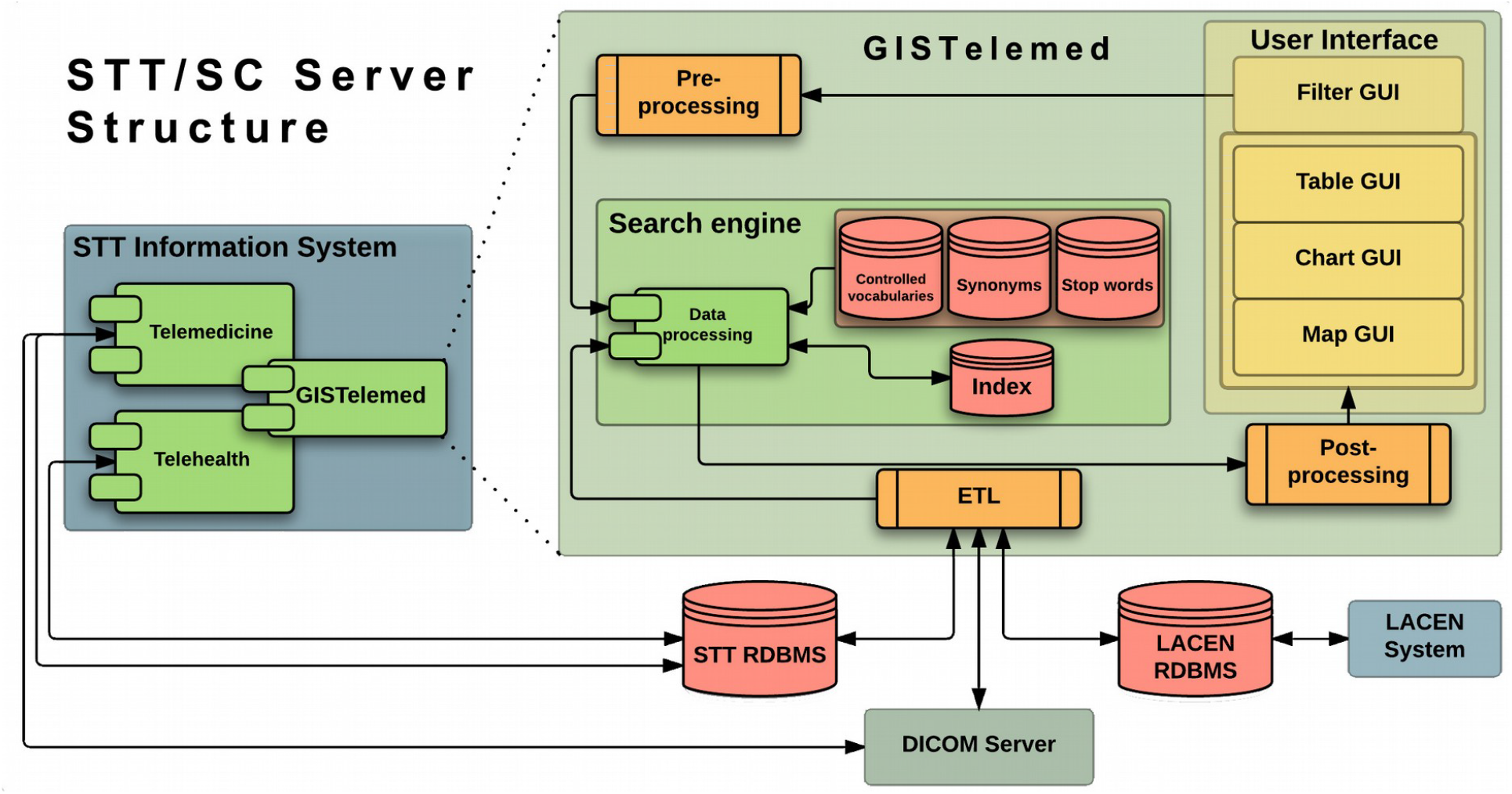
The GISTelemed architecture integrated into the context of STT/SC

The GUI (figure 2) is designed to support both experienced and inexperienced users in performing queries and epidemiological analyses. The filter tool (a) allows to select search parameters from a predefined set of filters based upon the controlled vocabularies employed for the indexation of the databases, as well as to use free text search. The interface also allows the user to choose which resulting data is to be displayed using a drag-and-drop mechanism.

**Figure 2.**
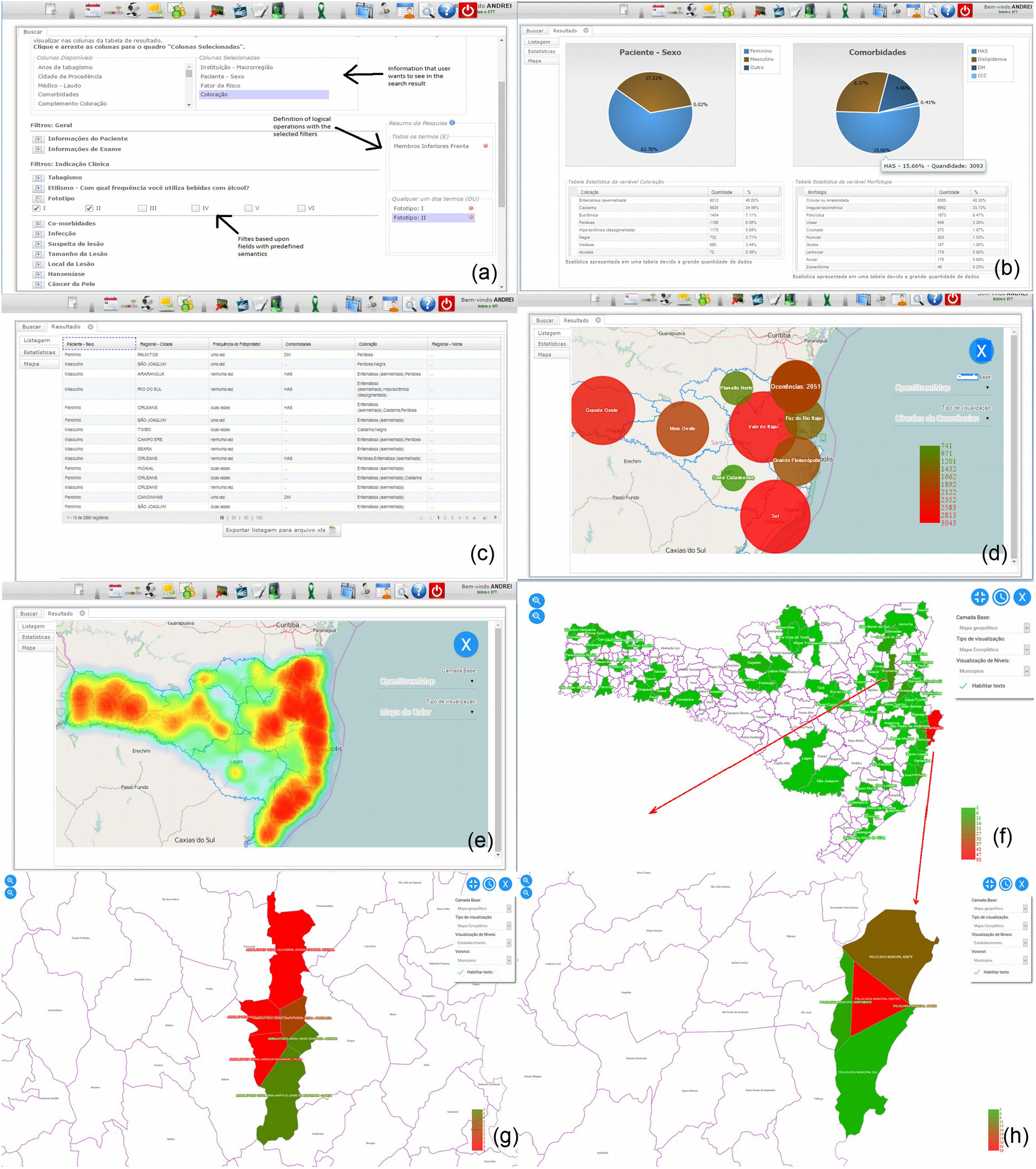
The GISTelemed user interface – search interface (a), charts (b) tables (c), and results in maps: occurrence circles (d), heat map (e), and as choroplethic map, showing municipalities (f) and voronoi diagrams depicting influence areas of individual healthcare facilities (g and h).

The graphic output tool displays the search results as charts (b), tables (c), and three types of maps: occurrences (d), heat maps (e), and a multiresolution choropletic representation that represents the results as areas of influence and can show the result per county, per municipality or per individual healthcare facilities, employing municipality borders and Voronoi diagrams as intra-municipal delimiters (f, g and h). The map of the Santa Catarina state is loaded using the WMS (Web Map Service) provided by SIG@SC, a service previously developed at our research group as a partnership between the Federal University of Santa Catarina (UFSC) and the Santa Catarina State Datacenter (CIASC) that offers public access to vector and raster geographic data. [27]

Another feature from GISTelemed is the timeline simulation: the user can take the results of a search and ask GISTelemed to present them in an animated timeline, where occurrences are shown in adjustable intervals and with a preajusted geographical granularity (county, municipality or healthcare facility). In Figure 3 the distribution of skin cancer cases in Santa Catarina between 2015 and 2018 was first calculated and then simulated month-wise, showing their municipalities of occurrence: figure 3(a) shows cases for January 2018 and figure 3(b) shows cases for August 2018. A slider automatically advances the timeline, producing an animated simulation. We provided additional video and image material on our site, at: http://site.telemedicina.ufsc.br/gistelemed/. The first video provided with this additional material shows several examples of this feature.

**Figure 3.**
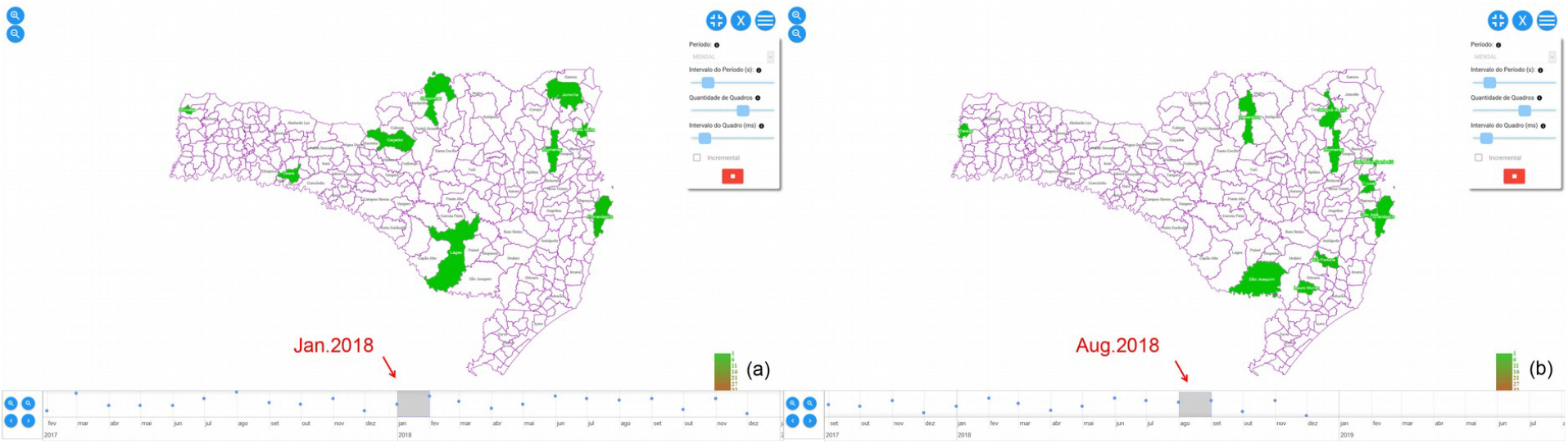
The GISTelemed timeline user interface showing two moments of a query on skin cancer occurrences.

### Evaluation strategy

We evaluated GISTelemed accordingly to the guidelines published by the US Centers for Disease Control and Prevention (CDC).[21] Due to its specificity, each attribute was assessed individually using criteria from the related literature, data from information systems available to this work, and input provided by a questionnaire customized according to AdEQUATE (questionnAire for Evaluation of QUAlity in TElemedicine systems). [22-25]

### Evaluation Dataset

We evaluated GISTelemed using:

- data from 62,393 tele-dermatoscopic examinations, covering cases of leprosy, skin cancer and psoriasis (23,271 cases) and other dermatoses (39,122 cases), performed between January/2015 and December/2017, employing the protocols defined in [28], and,
- responses from 39 healthcare professionals working in the Public Health area to the AdEQUATE questionnaire.

When data collection started (see Figure 4..a) there were 77 primary healthcare facilities in 73 municipalities of the State performing tele-dermatological examinations and triage. In December 2017 there were 313 facilities in 284 municipalities habilitated to perform tele-dermatological examinations. 16 of the new facilities implanted during the period were located in prisons. Most new facilities were implanted during 2015. During the data collection period, an average of 1,733.14 examinations/month was performed (see Figure 4..b).

**Figure 4.**
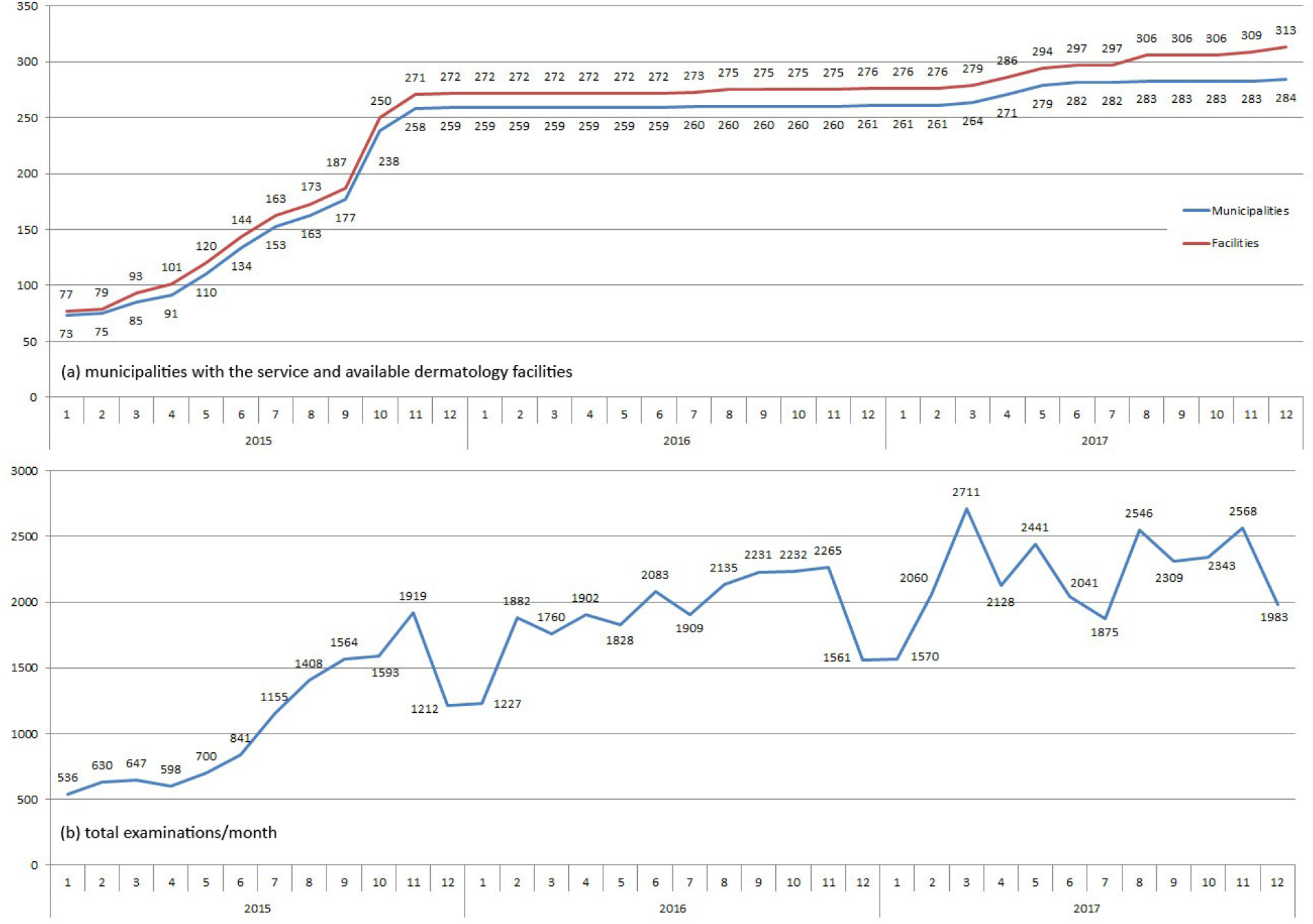
Sites performing tele-dermatological examinations in the State of Santa Catarina and number of tele-dermatoscopic examinations performed

### Gound Truth

As a ground truth to compare our epidemiological data against we chose the data on leprosy registered on the Brazilian National Information System on Reportable Diseases (SINAN).[7] Figure 5. shows a comparison between the 528 leprosy cases reported on SINAN and the 114 leprosy cases identified through tele-dermatology during the period 2015-2017.

**Figure 5.**
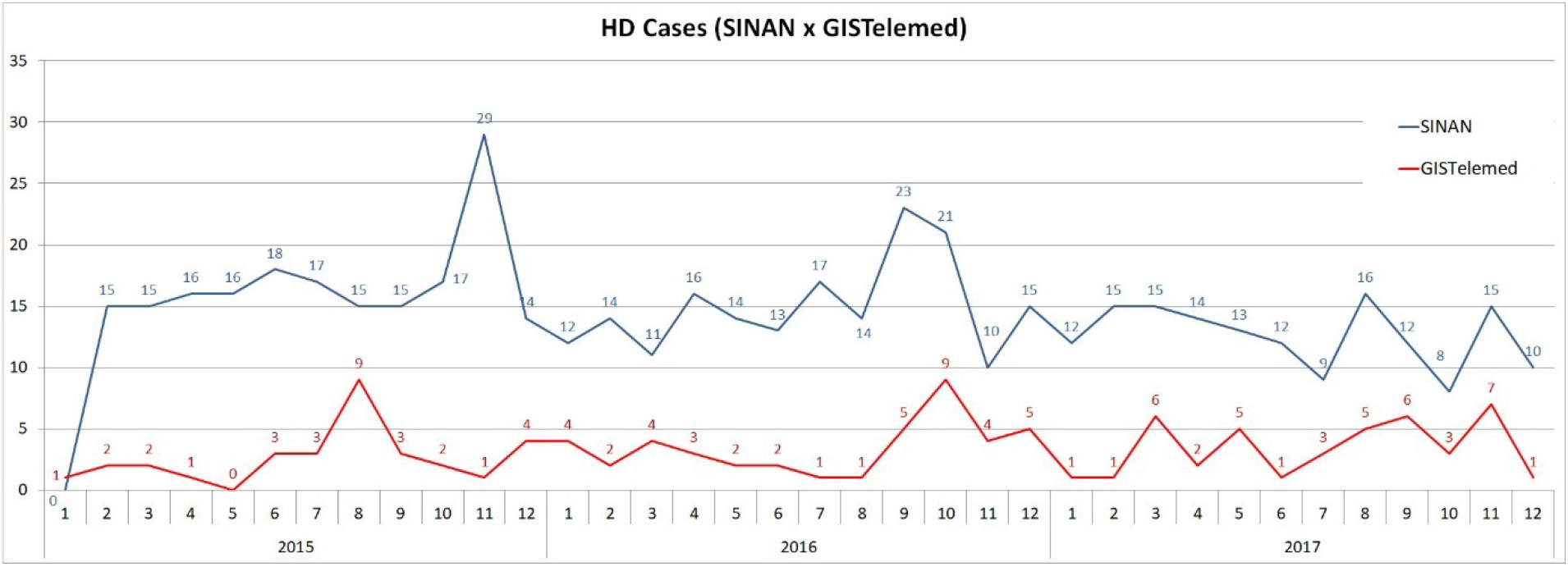
Comparison between leprosy cases reported on SINAN and those identified on GISTelemed

### Evaluation Parameters

In order to adhere to both [21] and [22-25], we evaluated GISTelemed according to the following criteria:

- <u>Usefulness</u> - degree to which a user is satisfied with its perceived achievement of pragmatic goals, including the contribution to the prevention and control of adverse health-related events or if it helps to determine that an adverse health-related event, previously thought to be unimportant, is actually important. This attribute was evaluated by the *Usefulness* subcharacteristic of the AdEQUATE model, assessing the degree to which a user is satisfied with its perceived achievement of pragmatic goals by using GISTelemed.
- <u>Simplicity</u> - structuredness and ease of operation of GISTelemed. This attribute was evaluated by the *Usability* characteristic of the AdEQUATE model, assessing the degree to which a product or system can be used by specific users.
- <u>Flexibility</u> - system’s adaptability to changes in the information needs or operating conditions. This attribute was evaluated by the *Flexibility* subcharacteristic of the AdEQUATE model, assessing the degree to which GISTelemed can be used in contexts beyond the ones initially specified.
- <u>Data quality</u> - completeness and validity of data stored by the system. We evaluated the completeness of patient and diseases data collected by GISTelemed, considering *excellent* data where the percentage of completeness is greater than 90%; *good* a percentage of completeness between 70.1% and 90%; and *poor* when the percentage of completeness is equal to or less than 70%.
- <u>Acceptability</u> - willingness of individuals and organizations to participate in using the system. Was measured by analyzing interview completion rates and question refusal rates, evaluated by the percentage of users who accepted to contribute with the study answering the available questionnaire.
- <u>Sensitivity</u> - proportion of all cases identified by the system that were also confirmed through and identifiable notification in SINAN to all cases retrieved by the system. This attribute was evaluated by comparing confirmed cases of leprosy registered both in GISTelemed and SINAN. Other diseases whose cases are registered in GISTelemed (skin cancer, psoriasis and other dermatoses) were excluded from this part of the evaluation due to the unavailability of a notification system for comparison. We employed the following formula:

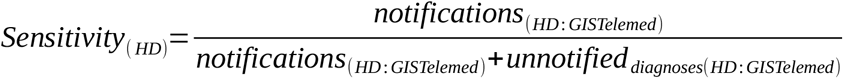
- <u>Positive Predictive Value (PPV)</u> - proportion of all leprosy cases notified in SINAN to the leprosy cases (Hansen’s disease = HD) notified in SINAN plus all retrieved cases in GISTelemed where leprosy was a diagnosis dermatoscopically confirmed by a medical report issued by a dermatologist but that were absent in the SINAN database. We employed the following formula:

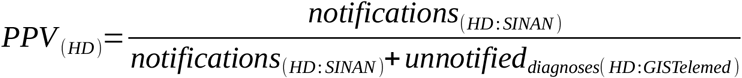
- <u>Representativeness</u> - degree to which the system accurately describes the occurrence of a health-related event over time and its distribution in the population by place and person. This attribute was also evaluated comparing leprosy cases in both GISTelemed and SINAN. For this purpose we employed a correllation analysis between the two curves and also a Bland-Altman plot analysis.
- <u>Stability</u> - ability of the system to collect, manage, and provide data satisfactorily, and to be operational and accessible when it is required for use. This attribute was evaluated by the *Reliability* characteristic of the AdEQUATE model, including its *Maturity, Availability, Fault tolerance* and *Recoverability* subcharacteristics, in order to assess the degree with which GISTelemed performs specific functions, under specific conditions, for a specific period of time.

### Software quality assessment

We invited users of STT/SC to follow a predefined script using GISTelemed to perform a set of search and retrieval tasks. Once the tasks were concluded, the participants filled the questionnaire intended to collect data on their perception of quality regarding GISTelemed. All questionnaire items were formulated as assertions, aiming to eliminate misinterpretation and to reduce the probability of error in responses. The responses follow a 4-point Likert scale, including the alternatives “Not applicable”, “Don’t know”, and “Don’t understand the item”, and were used to compute the distribution of each option for each questionnaire item, using a SUS-like approach for summarization.[29-30]

We analyzed the responses to the questionnaire using descriptive statistics, adopting the median as a representative for the middle of the distribution, and quartiles for grouping.[ 30] The score of responses given to each questionnaire item was calculated in a similar way of SUS scores,

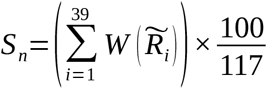

where 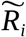, corresponds to the median of responses to the questionnaire for subcharacteristic *n* by respondent *i*, and 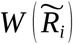 is a weight function based on a response option assuming the following values: 3 for “Totally agree”, 2 for “Agree”, 1 for “Disagree”, and 0 for “Totally disagree”. The response options “Don’t know”, “Not applicable”, and “Don’t understand the item” are ignored. The last component in the formula guarantees that *S*_n_ varies from 0 (poorest degree of quality − i.e. all respondents choose “Totally disagree”) to 100 (exceptional degree of quality − i.e. all respondents choose “Totally agree”).[31]

## RESULTS

From a total of 48 invitations, 39 healthcare professionals working in Epidemiological Vigilance and other Public Health-related areas from 13 municipalities of the Santa Catarina State accepted to participate on the assessment (a response rate of 81.2%). The distribution of the respondents by occupation is given in Figure 6..

**Figure 6.**
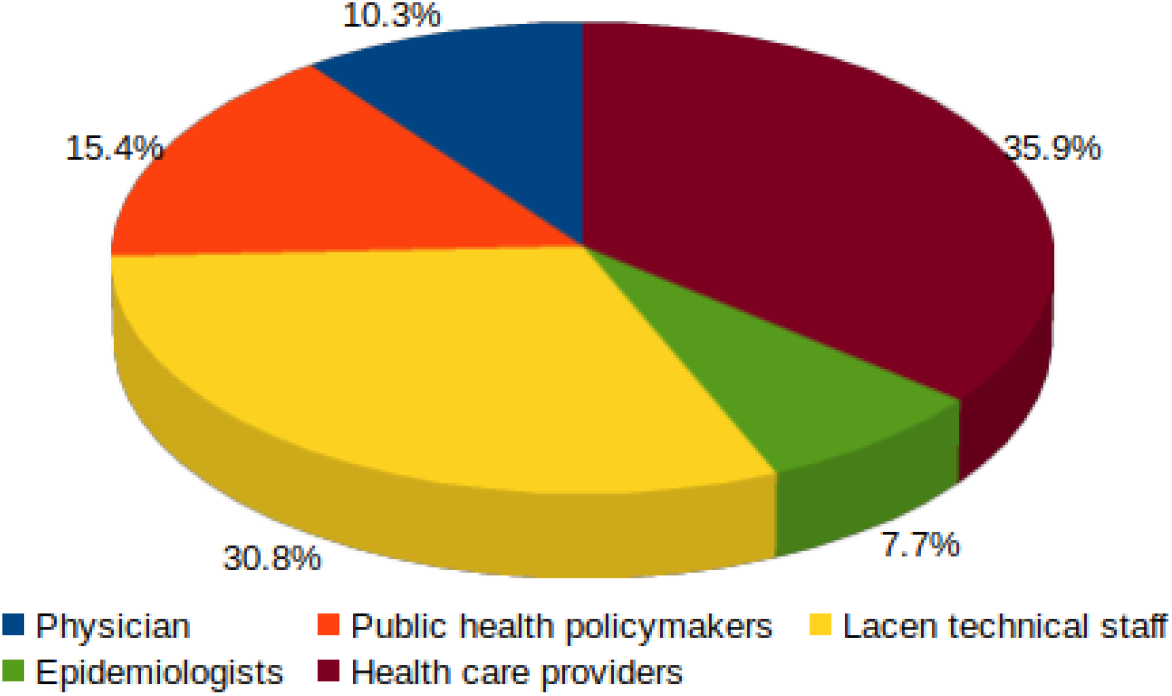
Distributions of respondents by occupation

All examinations were performed at primary healthcare facilities, where a local general practitioner performed the patient assessment and a trained technician performed the dermatoscopic examination. Data were uploaded using the telemedicine system and a tele-dermatologist analyzed the patients’ assessment report and the dermatoscopic examination and provided a triage report. We identified 114 leprosy cases with GISTelemed. Of these, 26 were cases we could not identify in SINAN, which we treated as unnotified cases, totaling 22.8% of all cases identified with GISTelemed, mostly from small upstate municipalities. A comparison of occurrences per municipality is shown in the Appendix Occurrences Monitored per Municipality.

### System Performance

From the point of view of data quality, search results with GISTelemed are shown in Table 1.: we identified a completion rate of 93.77% associated to the data provided by the primary health care facilities that first examined the patients.

**Table 1.**
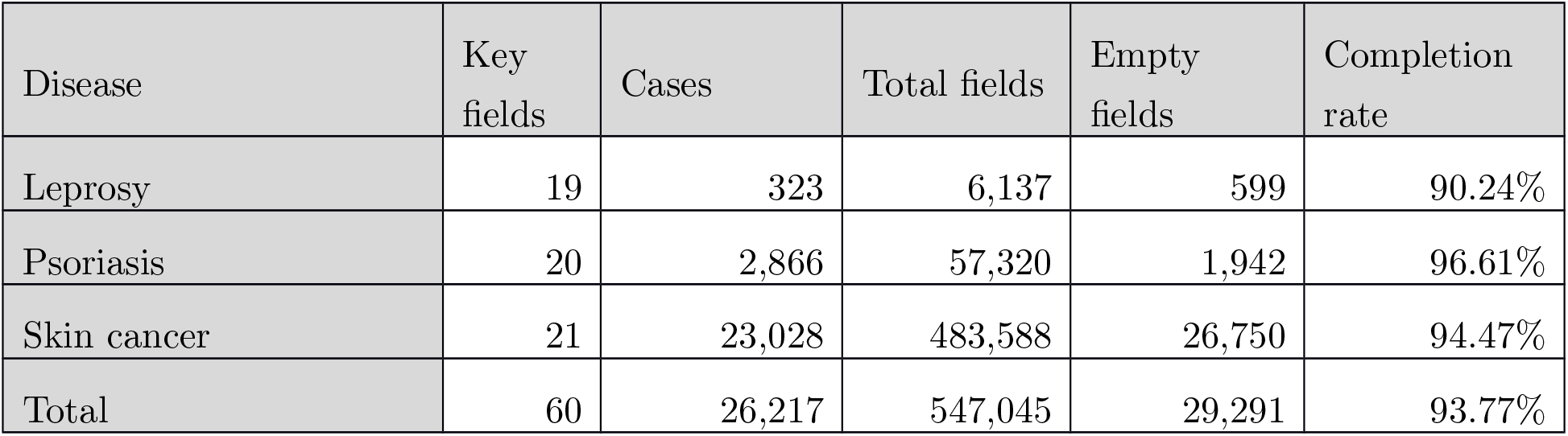
Data quality assessment by diseases

### Sensitivity, Positive Predictive Value and Representativeness

The sensitivity and PPV of GISTelemed for predicting notified leprosy cases were respectively 77.2% and 95.3%, when employing the formulas given above. From the point of view of representativeness, the correlation of the SINAN and GISTelemed leprosy curves shown in Figure 5. was 0.14348 for the whole 2015-2017 period. The correlations for individual years were respectively −0.01922 for 2015, 0.44495 for 2016 and 0.37384 for 2017. A Bland-Altman plot analysis showed a bias of 11.22 between both curves; with 95% confidence interval between 1.82 and 20.63. There were two outliers: January 2015, with a difference of −1 and November 2015, with a difference of 28. Figure 7. shows the comparison of SINAN and GISTelemed by a scatter plot (a) and a Bland-Altman plot (b).

**Figure 7.**
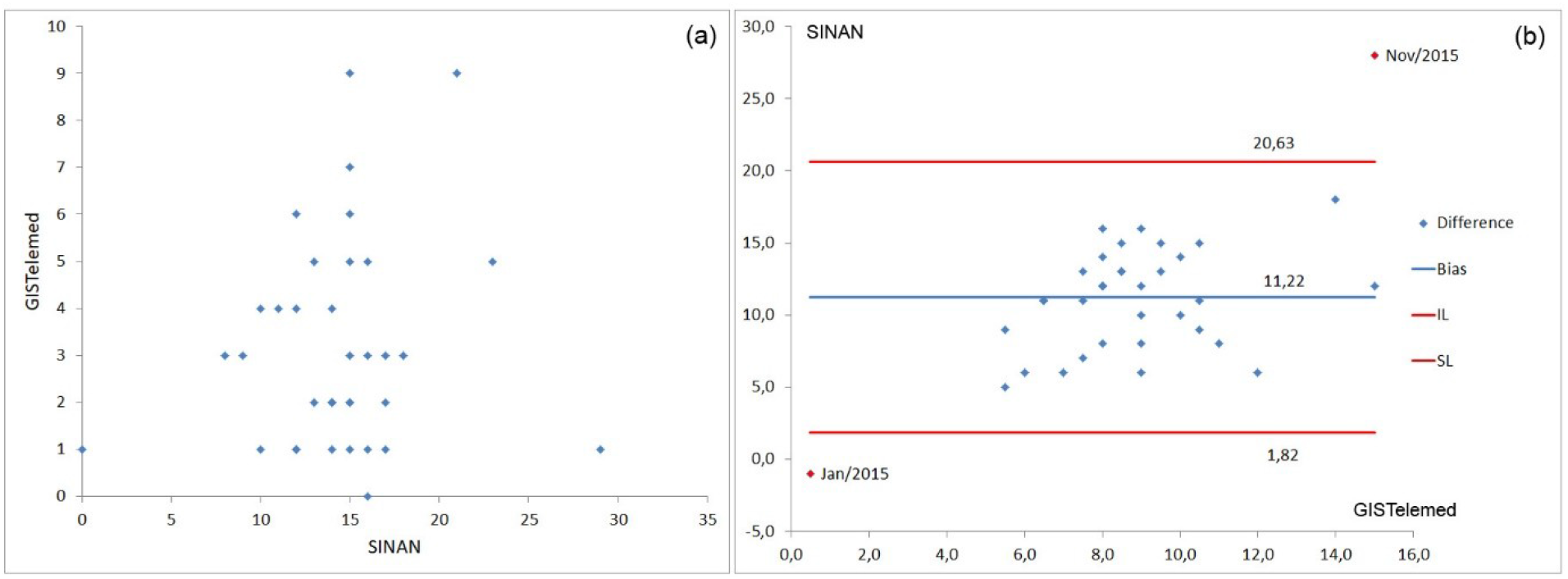
Scatter plot (a) and Bland-Altman plot (b) for SINAN and GISTelemed leprosy cases

### Software Quality

We analyzed the data acquired through the questionnaires using descriptive statistics, as described above. Scores for each subcharacteristic are presented in Figure 8., varying from 0 to 100. With a median over 50, we observed a good perception of the quality of GISTelemed. The interquartile range of 14.4 demonstrates low variability between groups.

**Figure 8.**
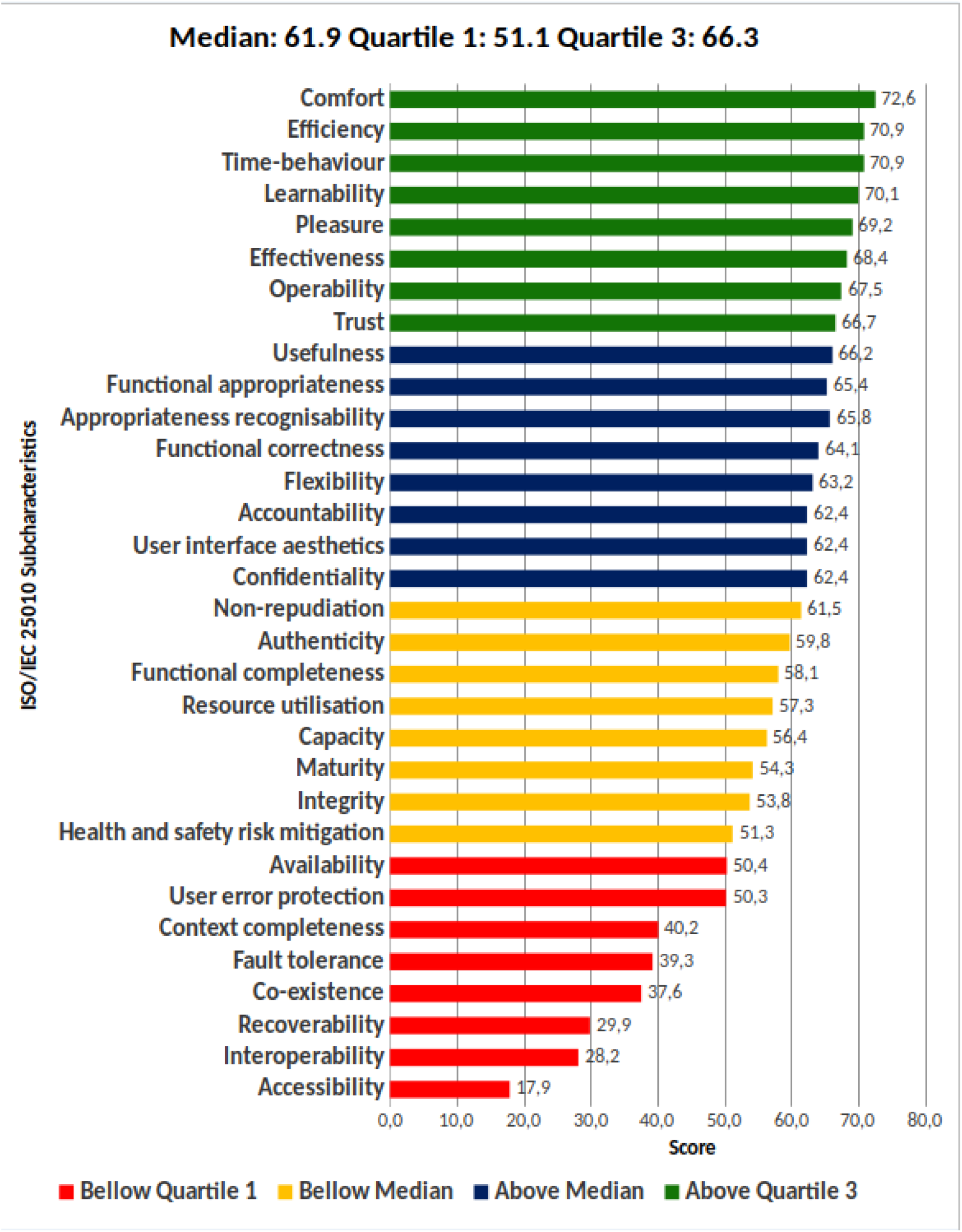
Scores for quality subcharacteristics of the ISO/IEC 25010 standard

Some subcharacteristics shown as “Bellow Quartile 1” received low scores because the responses “Not applicable”, “Don’t understand the item”, and “Don’t know” were not considered in the score calculation, especially the subcharacteristics with a score below 50 that have less than 60% of the responses scored positively or negatively. We can also observe in Figure 8. that, according to the users, the worst subcharacteristics are “Accessibility”, “Interoperability” and “Recoverability”. Nevertheless, since the median is over 50, a positive perception of the quality for GISTelemed can be noticed. Figure 9. presents the proportion of responses by the ISO/IEC 25010 standard subcharacteristics. We can observe differences in the position for some subcharacteristics when compared to Figure 8.; it occurs because the chart in Figure 9. shows the real distribution of answers, and it is presented in descending order, subcharacteristics evaluated positively first. “Time-behavior”, for instance, despite having the same score of “Efficiency” calculated by (1), has a better position due to its greater number of positive responses. We also observed that more than 50% of the users answered “Not applicable” or “Don’t know” for the subacharacteristics “Recoverability”, “Interoperability”, and “Accessibility”; it is an indication that users may had difficulty with these concepts, or that respondents lacked experience using GISTelemed. “Accessibility”, having more than 70% of its responses being “Not applicable”, may indicate an issue in GISTelemed regarding this subcharacteristic, or a bias in selecting respondents − from all participants, no one had disabilities.

**Figure 9.**
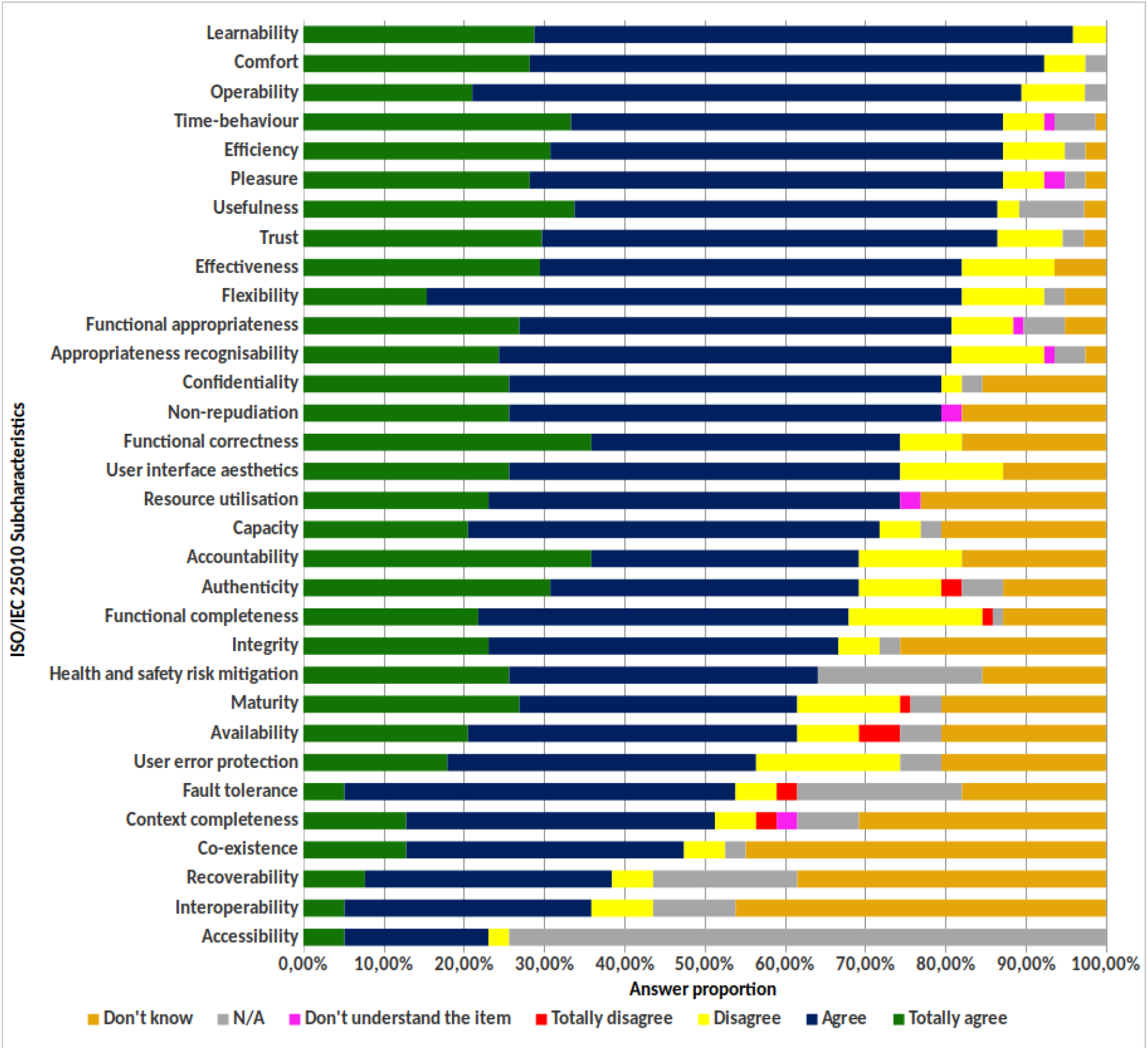
Proportion of responses by ISO/IEC 25010 subcharacteristics

**Figure 10.**
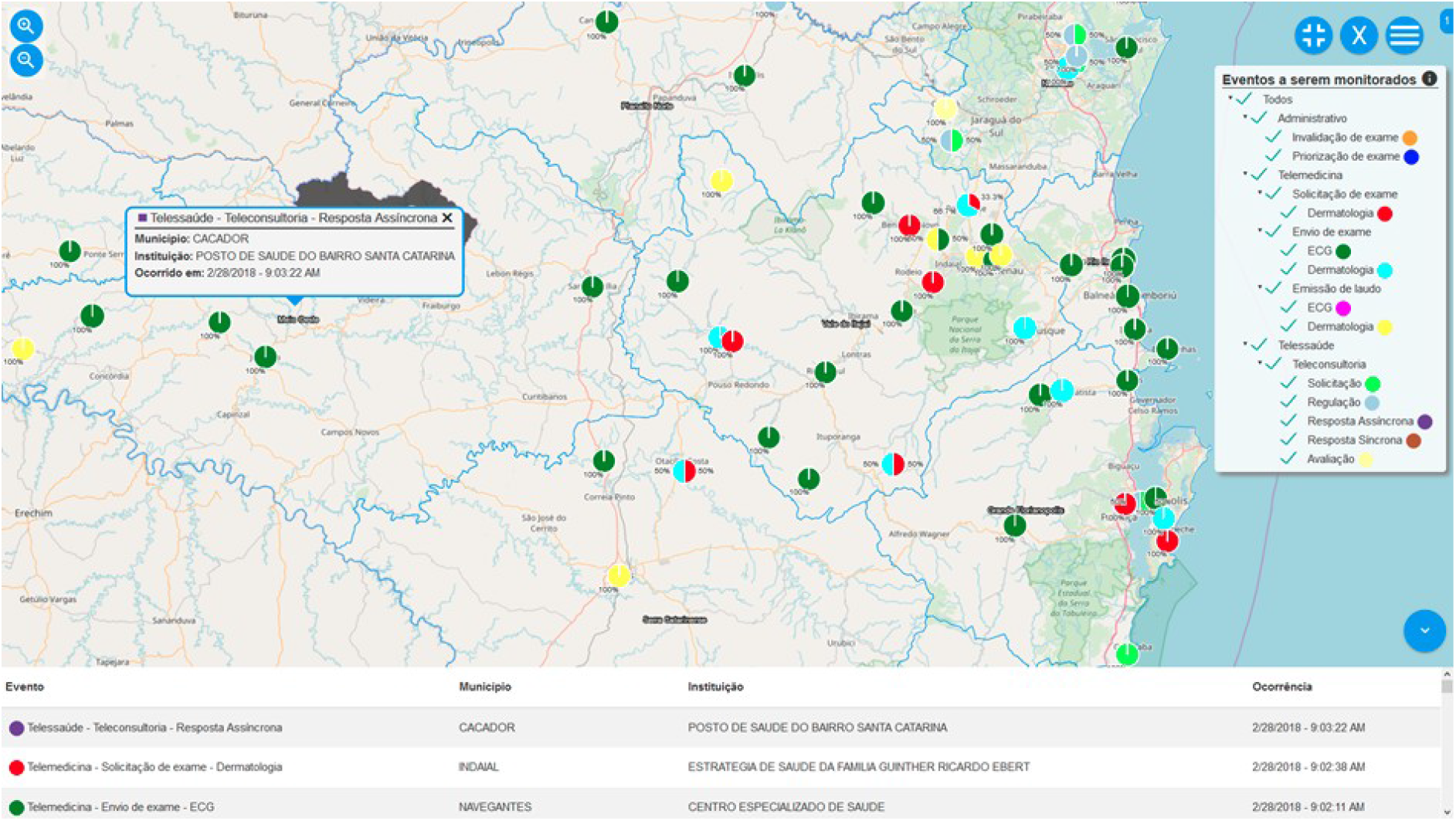
“Situtation room” real-time event monitoring module of GISTelemed

## DISCUSSION

We presented GISTelemed, a georeferenced epidemiological analysis tool integrated into a Telemedicine and Telehealth Information System. Intended to be used by medical staff, public health policymakers, and researchers, the module was developed to help on the study of chronic and infectious conditions, on guiding the healthcare decision-making; and on contributing to the development, monitoring and evaluation of interventionist actions for controlling and preventing diseases, providing fast feedback to public health policies. For the quantitative evaluation of GISTelemed, even if the STT/SC stores data from a broad spectrum of examinations, ranging from simples patient interviews up to magnetic resonance imaging (MRI) and computed tomography (CT) scans, we limited the scope to dermatology data because, for a part of these data (leprosy cases), we were able to compare our results with an external reference, which we took as our golden standard: the leprosy notifications stored on SINAN.

We evaluated the GISTelemed tool employing an approach that was as adherent to [21] as possible, given the limitations posed by the Brazilian environment where we tested it. This led us to consider it from two points of view: (a) *consistence with an external epidemiological data source*, where we took (leprosy epidemiological data from the Brazilian National Information System on Reportable Diseases (SINAN) [7] as a reference, and (b) *user perception*, where we employed an evaluation instrument from the field of Telemedicine [22]. We then interpreted the results we obtained accordingly to the guidelines given by [21].

### Consistency and Representativeness of Data

The analysis of the leprosy cases identified through GISTelemed, when compared to SINAN shows that GISTelemed identified much less cases. This was expected, since tele-dermatology is not the only point-of-entry for leprosy cases in the healthcare system as a whole. On the other side, correlation between data, even if positive but very poor for the whole period (0.14348), become much better for the periods of 2016 and 2017, when they are taken separately (0.44495 and 0.37384 respectively). According to the Rule of Thumb for interpreting the size of a correlation coefficient, values between 0.30 and 0.50 are classified with low positive correlation and a correlation between 0.00 and 0.30 is classified as negligible correlation [32]. We obtained similar results with the Bland-Altman plot: a bias of 11.22 shows that the results for SINAN are generally much higher, but the outliers in the curve comparison lay all in 2015. Both correlation analysis and Bland-Altman plot results between the SINAN and GISTelemed leprosy-curves are much better for the periods of 2016 and 2017, when tele-dermatoscopy coverage of the State was practically total. This reinforces an informal visual analysis of these curves, which shows that the curves in figure 4 tend to be similar in form in 2016 and 2017, even if the GISTelemed curve shows much lower amplitude. We understand that this indicates that in our State leprosy morbidity in telemedicine is reflecting a subset of the total morbidity, and that telemedicine-detected occurrences and overall occurrences tend to follow a similar case occurrence pattern. Since monitoring in telemedicine, being near-realtime, is much faster than in national notification systems such as SINAN, where notifications sometimes take months to become online, our results evidentiate that telemedicine-based epidemiology could be used as a very fast indicator of tendencies and changes in morbidity patterns and also help identify subit outbreaks. On the other side, GISTelemed identified 26 cases which were not reported in SINAN, mostly from very small municipalities. These could indicate a subnotification situation, which with the widespread use of telemedicine can now automatically be identified in near-realtime. They could also be old cases, which migrated from some private clinic to the public healthcare system with the new service offer represented by the public tele-dermatology service.

### User perception

Results from our case study show a good evaluation regarding the perceived software quality, according to measures encompassing “Comfort”, “Efficiency”, “Usability”, “Effectiveness”, “Operability”, “Performance efficiency”, and “Learnability”. Some evaluated items did not perform so well, such as “Accessibility”, “Interoperability”, “Recoverability”, and “Co-existence”. Considering, however, that most of the responses given for the questionnaire items were positive, the median calculated for all of the subcharacteristics was 61.9, and that 81.2% of the subcharacteristics have shown a score greater than 50, we can accept a good perception of quality, usefulness and acceptance of the GISTelemed module by the end users of the STT/SC information system.

Our perception was that the integration of spatial epidemiology tools to the STT/SC system, besides enabling visualization of data in maps, allowed users to analyze the evolution of morbidities and their co-occurrences in quasi-real time.

### Threats to validity

We identified the following threats to the validity of the conclusions arrived here:

1. Internal validity: There is a possible selection bias, because we invited only medical staff, municipal public health officers and public health researchers working directly with the STT/SC to respond the questionnaire, as they are the direct potential users. A broader study, involving a larger, external public has yet to be performed. Another threat related to a selection bias was that some respondents had their first contact with GISTelemed during the evaluation process, which may have influenced the answers for some items. To minimize this threat, the options “Not applicable”, “Don’t know”, and “Don’t understand the item” were included as valid answers.
2. Construction bias: The AdEQUATE evaluation model has not yet been extensively validated in other contexts; however, we believe that this threat is minimized because the adopted questionnaire had its items systematically derived from the ISO/IEC 25010 standard, TAM and SUS, which are largely used methods.

### New Features: Real-Time Monitoring of Events

The GISTelemed tool is an ongoing and evolving project and not all features could be evaluated in this study. In its more recent version, it also allows the real-time monitoring of events ocurring on the Telemedicine and the Telehealth networks and, more recently, all pacient referral requests *(tratamento fora de domicílio* - TFD), as shown in the figure below. We call this operation mode “Situation Room”. In its present version, this module offers a fixed pallete of events to be monitored, that can be selected to be watched, as shown in the figure. Events are organized per originating Primary Healthcare Facility. Last event is shown in a pop-up window. This module is intended for healthcare policies planning and for real-time monitoring of epidemiological processes.

We also provided additional time-lapse 10x speed video on our site, at: http://site.telemedicina.ufsc.br/gistelemed/ that illustrates this new module. The second video provided with this additional material shows 4 hours of operation of this feature.

### Conclusions and Future Work

This first edition of the GISTelemed module is still limited: it can research only cardiology, dermatology and clinical analyses (including infectious diseases) data and the capabilities of cross-referencing between these three data sources at an individual patient level are also poor, partially due to the lack of demographic integration of Brazilian public primary healthcare data. However, this first evaluation of GISTelemed has shown that an interactive spatial epidemiology tool can consistently be developed directly on top of a telemedicine infrastructure and be presented to the users as an integral part of this infrastructure. From both evaluation points of view, *consistency and representativeness of data* and *user perception* the module presented good results and has been considered a useful tool by healthcare personnel and policymakers in medium and small municipalities in Southern Brazil.

The utility of such a tool for healthcare policymakers of individual municipalities can be seen, e.g., on Figure 2.(g) and Figure 2.(h), where the different tendencies of the distribution of leprosy cases in a municipality can be seen: in Blumenau, Figure 2.(g), leprosy cases concentrate on the northwestern rural areas of the municipality, whereas on the Santa Catarina Island, Figure 2.(h), leprosy cases concentrate in downtown Florianópolis, suggesting different epidemiological mechanisms. The timeline visualization (Figure 3.) is also usefull to observe morbidities through time. In the video provided with this article, we show 3 different interactions. In the second interaction, we show cases of skin cancer on the tip of the nose through time: it becomes clear that the municipalities of Blumenau, Joinville (german settled areas with a predominantly fair-skinned population) and Florianópolis (fishing industry, watersports, and beaches) not only show a much higher prevalence but also show a steady number of occurences through time, whereas other regions are much more variable. This tool is new and there is nothing comparable being offered in the Brazilian Public Health area, our first validation study, however, has shown that local public health professionals found it useful and usable.

In 2018 we started to employ GISTelemed for the evaluation of the impact of the Santa Catarina State Telemedicine initiative and to perform cost analyses [33]. In 2019 we will start migrating all other areas of the STT/SC (teleradiology, obstetrics, tele-EEG) to DICOM Structured Reporting findings descriptions based upon controlled vocabularies. A new patient identification policy has also been introduced. This will allow us to index our data much better and also to broaden the search capabilities to these examination areas.

## Data Availability

All data referred to or described in the article are public.

http://site.telemedicina.ufsc.br/gistelemed/

## Acknowledgments

This work was supported by the SES/SC (Santa Catarina State Health Office), FAPESC (Fundação de Amparo à Pesquisa e Inovação do Estado de Santa Catarina) and IFSC (Instituto Federal de Santa Catarina).

## APPENDIX

Comparison of Leprosy Occurrences Monitored per Municipality

Comparison of Leprosy Occurrences Monitored per Municipality
| Municipality | GISTelemed | SINAN | Non-Notified |
| --- | --- | --- | --- |
| ABELARDO LUZ | 1 | 1 | 0 |
| ÁGUA DOCE |  | 4 | 0 |
| ÁGUAS MORNAS | 1 |  | 1 |
| ANCHIETA | 1 | 11 | 0 |
| ANGELINA | 1 |  | 1 |
| ARAQUARI |  | 1 | 0 |
| ARARANGUÁ |  | 1 | 0 |
| BALNEÁRIO BARRA DO SUL |  | 1 | 0 |
| BALNEÁRIO CAMBORIÚ |  | 12 | 0 |
| BALNEÁRIO PIÇARRAS |  | 3 | 0 |
| BARRA BONITA |  | 2 | 0 |
| BARRA VELHA |  | 2 | 0 |
| BELMONTE |  | 2 | 0 |
| BIGUAÇÚ | 1 | 2 | 0 |
| BLUMENAU | 5 | 14 | 0 |
| BOTUVERÁ |  | 2 | 0 |
| BRUSQUE | 3 | 12 | 0 |
| CAÇADOR | 1 | 8 | 0 |
| CAIBI |  | 2 | 0 |
| CAMBORIÚ |  | 6 | 0 |
| CAMPO ERÊ | 1 | 2 | 0 |
| CAMPOS NOVOS |  | 1 | 0 |
| CANELINHA | 4 | 1 | 3 |
| CANOINHAS | 2 | 7 | 0 |
| CATANDUVAS |  | 2 | 0 |
| CHAPECÓ | 3 | 29 | 0 |
| CONCÓRDIA | 2 | 15 | 0 |
| CRICIÚMA |  | 6 | 0 |
| CRISTOVÃO DO SUL |  | 1 | 0 |
| CUNHA PORÃ |  | 1 | 0 |
| DESCANSO |  | 1 | 0 |
| DIONÍSIO CERQUEIRA |  | 3 | 0 |
| FLOR DO SERTÃO | 1 | 2 | 0 |
| FLORIANÓPOLIS | 38 | 36 | 2 |
| FORQUILHINHA | 1 |  | 1 |
| FRAIBURGO | 2 | 6 | 0 |
| GAROPABA | 2 | 1 | 1 |
| GASPAR | 1 | 4 | 0 |
| GRÃO PARÁ | 1 |  | 1 |
| GUABIRUBA |  | 1 | 0 |
| GUARACIABA |  | 1 | 0 |
| GUARAMIRIM |  | 2 | 0 |
| GUATAMBU |  | 1 | 0 |
| HERVAL D'OESTE |  | 1 | 0 |
| IÇARA |  | 1 | 0 |
| ILHOTA |  | 2 | 0 |
| IMARUI | 1 |  | 1 |
| IMBITUBA | 2 | 1 | 1 |
| IMBUIA | 1 |  | 1 |
| INDAIAL |  | 1 | 0 |
| ITÁ | 1 |  | 1 |
| ITAIÓPOLIS |  | 1 | 0 |
| ITAJAÍ |  | 15 | 0 |
| ITAPEMA |  | 9 | 0 |
| ITAPIRANGA |  | 1 | 0 |
| ITAPOÁ |  | 1 | 0 |
| JAGUARUNA |  | 1 | 0 |
| JARAGUÁ DO SUL | 3 | 13 | 0 |
| JOINVILLE |  | 71 | 0 |
| LAGES | 1 | 9 | 0 |
| LAGUNA | 2 | 2 | 0 |
| LAURO MULLER |  | 1 | 0 |
| LEBON RÉGIS | 3 | 2 | 1 |
| MACIEIRA |  | 1 | 0 |
| MARAVILHA |  | 15 | 0 |
| MODELO |  | 1 | 0 |
| MONDAÍ |  | 2 | 0 |
| MONTE CARLO |  | 2 | 0 |
| MONTE CASTELO |  | 1 | 0 |
| MORRO GRANDE |  | 1 | 0 |
| NAVEGANTES |  | 11 | 0 |
| NOVO HORIZONTE |  | 1 | 0 |
| OURO |  | 1 | 0 |
| PAIAL | 1 |  | 1 |
| PALHOÇA | 4 | 5 | 0 |
| PALMA SOLA | 3 | 2 | 1 |
| PALMEIRA | 1 |  | 1 |
| PALMITOS |  | 1 | 0 |
| PAPANDUVA |  | 3 | 0 |
| PARAÍSO |  | 2 | 0 |
| PAULO LOPES | 1 |  | 1 |
| PENHA | 1 | 5 | 0 |
| PINHALZINHO | 1 | 2 | 0 |
| PINHEIRO PRETO |  | 1 | 0 |
| POMERODE |  | 3 | 0 |
| PONTE SERRADA |  | 1 | 0 |
| PORTO BELO |  | 1 | 0 |
| PORTO UNIÃO |  | 7 | 0 |
| PRAIA GRANDE |  | 2 | 0 |
| PRINCESA |  | 3 | 0 |
| QUILOMBO |  | 2 | 0 |
| RIO DO SUL |  | 9 | 0 |
| RIO NEGRINHO |  | 1 | 0 |
| RIQUEZA |  | 2 | 0 |
| ROMELÂNDIA |  | 5 | 0 |
| SALTO VELOSO |  | 1 | 0 |
| SANTO AMARO DA<br>IMPERATRIZ |  | 2 | 0 |
| SÃO BENTO DO SUL | 1 | 2 | 0 |
| SÃO BERNARDINO | 1 |  | 1 |
| SÃO CARLOS |  | 8 | 0 |
| SÃO DOMINGOS |  | 1 | 0 |
| SÃO FRANCISCO DO SUL |  | 4 | 0 |
| SÃO JOÃO BATISTA |  | 1 | 0 |
| SÃO JOAQUIM |  | 2 | 0 |
| SÃO JOSÉ |  | 13 | 0 |
| SÃO JOSÉ DO CEDRO |  | 4 | 0 |
| SÃO JOSÉ DO CERRITO | 1 | 1 | 0 |
| SÃO LOURENÇO DO OESTE | 1 | 1 | 0 |
| SÃO MIGUEL DA BOA VISTA |  | 2 | 0 |
| SÃO MIGUEL DO OESTE |  | 6 | 0 |
| SÃO PEDRO DE ALCÂNTARA | 1 | 2 | 0 |
| SAUDADES |  | 1 | 0 |
| SCHROEDER |  | 1 | 0 |
| SEARA | 1 |  | 1 |
| SIDERÓPOLIS |  | 1 | 0 |
| SOMBRIO |  | 1 | 0 |
| SUL BRASIL |  | 1 | 0 |
| TIJUCAS | 1 |  | 1 |
| TIMBÓ | 2 | 3 | 0 |
| TRÊS BARRAS |  | 7 | 0 |
| TREZE DE MAIO | 1 |  | 1 |
| TUBARÃO |  | 6 | 0 |
| TUNÁPOLIS | 1 | 1 | 0 |
| UNIÃO DO OESTE |  | 2 | 0 |
| VIDAL RAMOS |  | 1 | 0 |
| VIDEIRA | 1 | 11 | 0 |
| XANXERÊ |  | 4 | 0 |
| XAXIM | 1 | 4 | 0 |
| DIONISÍO CERQUEIRA | 1 |  | 1 |
| JOAÇABA | 2 |  | 2 |
| **Totals** | **114** | **528** | **26** |

## REFERENCES

1. World Health Organization – Global Health Observatory (GHO), Brazil: country profiles. http://www.who.int/gho/countries/bra/country_profiles/en/ (accessed 10 Dec 2017).

2. Elliott P, Wartenberg D. Spatial epidemiology: current approaches and future challenges. Environ Health Perspect. 2004 Jun;112(9):998–1006. DOI: 10.1289/ehp.6735

3. Hemingway, H. et. al. Big data from electronic health records for early and late translational cardiovascular research: challenges and potential, European Heart Journal, ehx487, DOI: 10.1093/eurheartj/ehx487.

4. L.E. Thornton, J.R. Pearce, A.M. Kavanagh. Using Geographic Information Systems (GIS) to assess the role of the built environment in influencing obesity: a glossary. Int J Behav Nutr Phys Act, 8 (1) (2011), p. 1. DOI: 10.1017/CBO9781139003643.002.

5. Barreto SM, Miranda JJ, Figueroa JP, et al. Epidemiology in Latin America and the Caribbean: current situation and challenges. Int J Epidemiol 2012;41(2);557–71.

6. Fares RC, Souza KP, Añez G, et al. Epidemiological Scenario of Dengue in Brazil. Biomed Res Int 2015;2015;1–13.

7. Coelho, F.C. et al. Epidemiological data accessibility in Brazil. The Lancet Infectious Diseases, Volume 16, Issue 5, 524–525, May 2016.

8. Oliveira J.C.F., Leão A.M.M., Britto F.V.S. Analysis of the epidemiological profile of leprosy at Maricá, Rio de Janeiro: a contribution from nursing. UERJ Nursing Journal 2014;22(6). DOI:10.12957/reuerj.2014.13400.

9. Brazilian Health Ministry, Pan-American Health Organization and FundaÇão Oswaldo Cruz. The Brazilian Experience in Health Information Systems - Volume 1. http://bvsms.saude.gov.br/bvs/publicacoes/experiencia_brasileira_sistemas_saude_volume1.pdf (accessed 10 Dec 2017)

10. Groft S.C., de la Paz M.P. Rare diseases - avoiding misperceptions and establishing realities: the need for reliable epidemiological data. Adv Exp Med Biol 2010;686;3–14.

11. Mattos JSC, Mauad EC, Syrjänen K, et al. The Impact of Breast Cancer Screening Among Younger Women in the Barretos Region, Brazil. Anticancer Res 2013;33(6);2651–5

12. de Ávila AL, Krepischi AC, Moredo LF, et al. Germline CDKN2A mutations in Brazilian patients of hereditary cutaneous melanoma. Fam Cancer 2014;13(4);645—9.

13. Blozik E, Grandchamp C, von Overbeck J. Influenza surveillance using data from a telemedicine centre. Int J Public Health 2012;57(2);447–52.

14. Finet P, Le Bouquin Jeannès R, Dameron O, et al. Review of current telemedicine applications for chronic diseases. Toward a more integrated system? Innovation and Research in BioMedical engineering 2015;36(3);133–57.

15. Parmar P, Mackie D, Varghese S, et al. Use of telemedicine technologies in the management of infectious diseases: a review. Clin Infect Dis 2015;60(7);1084–94.

16. World Health Organization - Telemedicine: Opportunities and developments in Member States - Report on the second global survey on eHealth. http://www.who.int/goe/publications/goe_telemedicine_2010.pdf (accessed 10 Dec 2017).

17. Ohannessian R. Telemedicine: Potential applications in epidemic situations. European Research in Telemedicine 2015;4(3);95–8.

18. Inácio AS, Macedo DDJ, Andrade R, et al. Designing an information retrieval system for the STT/SC. IEEE 16th International Conference on e-Health Networking, Applications and Services. 2014;500–5.

19. Maia RS, Wangenheim Av, Nobre LF. A statewide telemedicine network for public health in Brazil. IEEE 19th International Symposium on Computer-Based Medical Systems. 2006;495–500.

20. Inácio AS, Macedo DDJ, Andrade R, et al. An architecture for information retrieval in a telemedicine system. IEEE 27th International Symposium on Computer-Based Medical Systems. 2014;513–4.

21. Centers for Disease Control and Prevention - Framework for Program Evaluation in Public Health. https://www.cdc.gov/mmwr/preview/mmwrhtml/rr4811a1.htm (accessed 28 Jan 2018).

22. Alves JM, Wangenheim CGv, Lacerda TC, et al. AdEQUATE Software Quality Evaluation Model v1.0. Relatórios Técnicos do INCoD 2015 Jan-Dec;5(1). http://www.incod.ufsc.br/wp-content/uploads/2015/12/Relatorio-Tecnico-AdEQUATE-v2.4a.pdf (accessed 28 Jan 2018).

23. International Organization for Standardization. ISO/IEC 25010:2011 - Systems and software engineering -- Systems and software Quality Requirements and Evaluation (SQuaRE) -- System and software quality models. http://www.iso.org/iso/catalogue_detail.htm?csnumber=35733 (accessed 14 Feb 2018).

24. Davis FD. Perceived usefulness, perceived ease of use, and user acceptance of information technology. MIS Q 1989;13(3);319–40.

25. Brooke J. SUS: A quick and dirty usability scale. Usability Evaluation In Industry 1996;189(194);4–7

26. Russell S. Kirby, Eric Delmelle, Jan M. Eberth, Advances in spatial epidemiology and geographic information systems, Annals of Epidemiology, Volume 27, Issue 1, 2017, Pages 1–9, DOI: 10.1016/j.annepidem.2016.12.001.

27. Santa Catarina State Government. Sistema de Informações Geográficas de Santa Catarina (SIGSC). http://sigsc.sc.gov.br/ (accessed 14 Apr 2018).

28. Piccoli M.F., Amorim B.D., Wagner H.M., Nunes D.H. Teledermatology protocol for screening of skin cancer. An Bras Dermatol. 2015;90(2):202–10. DOI: 10.1590/abd1806-4841.20153163

29. Sauro J, Lewis JR. When designing usability questionnaires, does it hurt to be positive? SIGCHI Conference on Human Factors in Computing Systems. 2011;2215–24.

30. Boone HN, Boone DA. Analyzing likert data. Journal of Extension 2012;50(2). https://www.joe.org/joe/2012april/tt2.php (accessed 18 Feb 2018).

31. Finstad K. The Usability Metric for User Experience. Interact Comput 2010;22(5);323–7.

32. Hinkle DE, Wiersma W, Jurs SG. Applied Statistics for the Behavioral Sciences. 5th ed. Boston: Houghton Mifflin; 2003.

33. von Wangenheim, A., Nunes, D.H. Direct Impact on Costs of the TeledermatologyCentered Patient Triage in the State of Santa Catarina - Analysis of the 2014–2018 Data. Technical Report INCoD/TELEMED.04.2018.E.01, ISSN 2236-5281, July, 2018. DOI: 10.13140/RG.2.2.20044.92807.

